# A Method for Sensitivity Analysis of Automatic Contouring Algorithms Across Different MRI Contrast Weightings Using SyntheticMR

**DOI:** 10.1101/2025.01.10.25319895

**Authors:** Lucas McCullum, Zayne Belal, Warren Floyd, Alaa Mohamed Shawky Ali, Natalie West, Samuel Mulder, Yao Ding, Jiaofeng Xu, Dan Thill, Nicolette O’Connell, Joseph Stancanello, Kareem A. Wahid, David T. Fuentes, Ken-Pin Hwang, Clifton D. Fuller

## Abstract

**Background:** Currently, a majority of institution-specific automatic MRI-based contouring algorithms are trained, tested, and validated on one contrast weighting (i.e., T2-weighted), however their actual performance within this contrast weighting (i.e., across different repetition times, TR, and echo times, TE) is under-investigated and poorly understood. As a result, external institutions with different scan protocols for the same contrast weighting may experience sub-optimal performance.

**Purpose:** The purpose of this study was to develop a method to evaluate the robustness of automatic contouring algorithms to varying MRI contrast weightings.

**Methods:** One healthy volunteer and one patient was scanned using SyntheticMR on the MR-Simulation device. The parotid and submandibular glands in these subjects were contoured using an automatic contouring algorithm trained on T2-weighted MRIs. For ground truth manual contours, two radiation oncology residents and one pre-resident physician were recruited and their STAPLE consensus was determined. A total of 216 different MRI TR and TE combinations were simulated across T1-, T2-, and PD-weighted contrast ranges using SyntheticMR’s post-processing software, SyMRI. Comparisons between automatic contouring algorithm contours and the ground truth were determined using the Dice similarity coefficient (DSC) and 95^th^ percentile Hausdorff distance (HD95).

**Results:** Notable differences in the automatic contouring model’s performance were seen across the contrast-weighted range, even within the T2-weighted range. Further, some models even performed as well or better across subsets of the T1-weighted range. The PD-weighted range saw the worst performance. The range of discrepancy in DSC and HD95 exceeded 0.2 and 3.66 mm, respectively, in some structures. In the T2-weighted contrast region where the model was trained, 100%, 40%, 24%, and 57% for the DSC in the left parotid, right parotid, left submandibular, and right submandibular gland, respectively, exceeded interobserver variability.

**Conclusions:** This study demonstrates the variable performance of MRI-based automatic contouring algorithms across varying TR and TE combinations. This methodology could be applied in future studies as a method for evaluating model sensitivity, out of distribution detection ability, and performance drift.

## 1. Introduction

Automated approaches have expanded in the field of medical physics and radiation oncology in recent years^1^. One of the primary explorations of their potential clinical utility is in automatic contouring of relevant structures for radiation therapy planning. Although super-human performance has been seen in several studies using traditional metrics such as the Dice similarity coefficient (DSC)^2,3^, there is still a large gap between the successful development and performance of these algorithms and their eventual clinical translation with a vast majority failing to cross this translational gap^4^.

A key step in validating model performance is quantifying their out of distribution (OOD) detection capabilities under the heterogenous conditions common in clinical practice. Taking it a step further, knowing how the model performs on OOD data, or data that the model has not seen during training, will help to expand its deployment across novel settings using a transfer learning approach without requiring the extra time needed to retrain a model from scratch. One study assessed the impact of slice thickness, pixel size, and computed tomography (CT) dose on the performance of automatic contouring algorithms on head and neck cancer patients and showed that significant deviations from baseline training data resulted in reduced model performance according to the DSC, Hausdorff distance (HD), and mean surface distance (MSD)^5^. Recent work investigating the metrics used to evaluate the performance of automatic contouring for radiation therapy treatment planning has suggested that multi-domain evaluation is essential to judge their clinical readiness^6^. The same paper suggested that there is no best metric to assess this, therefore multiple metrics must be combined for effective evaluation. However, other factors must be considered when effectively evaluating automatic contouring models such as their expected application space.

A previous study investigated the performance of various automatic contouring approaches for head and neck organs-at-risk (OARs) indicating that deep learning methods were the best tradeoff between performance and execution time^7^. However, deep learning models often require large, highly curated, datasets to effectively learn segmentation tasks^8^. This raises the problem of heterogenous datasets due to the desire to obtain more training data. In MRI, this can arise as different acquisition parameters (i.e., repetition time, TR, and echo time, TE) although, on the surface, each image will be labeled as T2-weighted, for example. The subtle issue here is that changing the TR will affect the amount of T1 signal expression while changing the TE will affect the amount of T2 signal nulling, thus generating different contrasts across different TR and TE parameter combinations^9,10^. Although, theoretically, enhancing the diversity of the training data should enhance the model performance^11^, this has not been tested rigorously across varying TR and TE in MRI data. Additionally, even across single institutions, a heterogenous distribution of T2-weighted images across different TR and TE is not uncommon and highly studied^12–14^ and can thus lead to uncertain model performance upon deployment.

Accordingly, to validate the performance of these models against varying clinically possible acquisition parameters, it would be beneficial to perform a sensitivity analysis of the model against a wide range of MRI acquisition parameters, even with the same contrast weighting (i.e., T2-weighting), using several clinically accepted metrics of automatic contouring algorithm performance. However, acquiring multiple MRI acquisition parameters across varying contrast weighted images on the same subject is time-intensive^15^, resource-intensive, and potentially unsafe due to the concerns of specific energy dose, or SED^16^. One recent innovation in MRI enabling synthetic contrast weighting is SyntheticMR, developed by the company SyntheticMR AB (Linköping, Sweden)^17^. Specifically, their post-processing package SyMRI can fit quantitative relaxometry maps and use that information to synthetically generate inherently co-registered contrast weighted images at varying TR and TE combinations as well as the addition of one or multiple inversion recovery pulses^18^. Further, a recent paper has demonstrated the feasibility of this technique in the radiation oncology workflow on both MR-Simulation and MR-Linac scanners^19^, opening the door for expanded investigation and its use in clinical practice.

In head and neck cancers, salivary glands are of critical importance to contour accurately due to their radiation sensitivity^20^ and direct association to the high prevalence of xerostomia, or dry mouth due to reduced salivary flow, as a result of radiation-induced damage^21^. This toxicity has been cited by patients as the most debilitating toxicity as a result of radiation therapy^22^ due to its persistence lasting greater than a year following treatment^23^ as well as its impact on social life and dental health^24^. Therefore, in this investigation we studied the performance of an automatic contouring model trained on T2-weighted MRI images applied specifically to the salivary glands in the head and neck. We evaluated the model’s performance sensitivity to varying TR and TE combinations across the T2-weighted contrast region as well as outside the model’s training data to include the T1-weighted and PD-weighted contrast regions as well.

## 2. Methods

### 2.1. MRI Imaging Data Acquisition

All MRI scans were performed on a 3T Siemens Vida MR-Simulation scanner (Siemens Healthcare; Erlangen, Germany). For this study, the SyntheticMR (Linköping, Sweden) 2D-multi-delay multi-echo (MDME) sequence was acquired with acquisition parameters as shown in Table 1. Further, due to the non-isotropic acquisition of 2D-MDME, all scans were acquired in the axial / transverse plane to best visualize the salivary glands in the head and neck^25^.

**Table 1:**
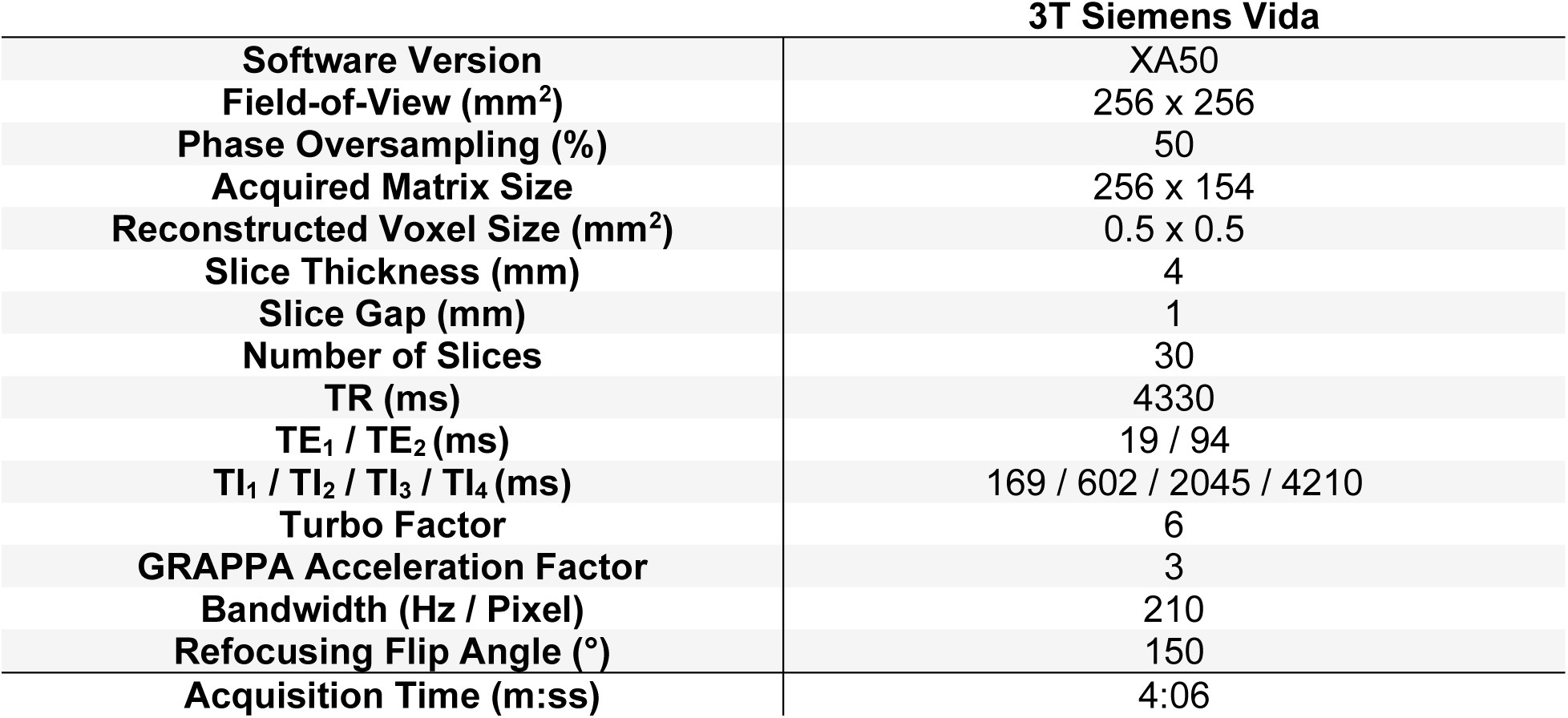
MRI acquisition parameters utilized in this study. Note, TI = inversion time.

| **3T Siemens Vida** |  |
| --- | --- |
| **Software Version** | XA50 |
| **Field-of-View (mm<sup>2</sup>)** | 256 x 256 |
| **Phase Oversampling (%)** | 50 |
| **Acquired Matrix Size** | 256 x 154 |
| **Reconstructed Voxel Size (mm<sup>2</sup>)** | 0.5 x 0.5 |
| **Slice Thickness (mm)** | 4 |
| **Slice Gap (mm)** | 1 |
| **Number of Slices** | 30 |
| **TR (ms)** | 4330 |
| **TE<sub>1</sub> / TE<sub>2</sub> (ms)** | 19 / 94 |
| **TI<sub>1</sub> / TI<sub>2</sub> / TI<sub>3</sub> / TI<sub>4</sub> (ms)** | 169 / 602 / 2045 / 4210 |
| **Turbo Factor** | 6 |
| **GRAPPA Acceleration Factor** | 3 |
| **Bandwidth (Hz / Pixel)** | 210 |
| **Refocusing Flip Angle (°)** | 150 |
| **Acquisition Time (m:ss)** | 4:06 |

### 2.2. Patient Description

A female healthy volunteer in their 20s and a male patient in their 50s with American Joint Committee on Cancer (AJCC) 8^th^ Edition Stage I (cT2, cN0, cM0, p16+) human papilloma virus (HPV) positive of oropharynx, squamous cell carcinoma of the left tonsil was scanned once on the MR-Simulation scanner using the SyntheticMR 2D-MDME sequence. All participants provided written informed consent and were consented to an internal imaging protocol (PA15-0418), both approved by the institutional review board at The University of Texas MD Anderson Cancer Center.

### 2.3. Data Collection and Image Processing

For the *in vivo* analysis, the parotid and submandibular glands were chosen for analysis as the primary OARs for salivary dysfunction and were contoured automatically using one automatic contouring algorithm in the Advanced Medical Imaging Research Engine (ADMIRE) research software (v3.48.4, Elekta AB, Stockholm, Sweden). The automatic contouring algorithm used a 3D-ResUNet^26^ deep learning approach with training on a unique patient atlas. A total of 44 paired MRI images with labels were used during training, 25 of which were 5 sets of 5 images from the same patient across different treatments on the MR-Linac device. The remaining images were from unique patients. Each MRI image had a spatial resolution of 1.8 x 1.8 x 1.2 mm^3^ with a matrix size of 224 x 224 x 64 with a T2-weighting with acquisition parameters of TR = 1535 ms, effective TE = 278 ms, and equivalent TE = 212 ms as further described in McDonald et al., 2024^7^. The input images for each model were pre-processed by thresholding to pixel values between the 0.25 and 99.75 percentiles for outlier removal, normalizing using the Z-score method, and scaling to between –1 and 1 using a linear transformation. The training was set to a fix number of 200 epochs where the first half of epochs used the same learning rate and the second half began to linearly decrease. Early stopping was not chosen because an online elastic deformation was applied on each patient to deform the image and masks on the fly to suppress potential overfitting. As a result, the final training accuracy averaged above 98.5% across the desired structures ignoring background.

The acquired 2D-MDME images were processed using SyntheticMR’s post-processing software, SyMRI (StandAlone 11.3.11, Linköping, Sweden). The same structures were contoured by two radiation oncology residents and one pre-resident physician in RayStation Research 12A R v13.1.100.0 (RaySearch Laboratories, Stockholm, Sweden) using a synthetic TR of 5254 ms and TE of 145 ms. This set of acquisition parameters was chosen as a representation of a general strongly T2-weighted image. The consensus ground truth contour for each structure was created using the simultaneous truth and performance level estimation (STAPLE) algorithm^27^ on all of the human contours (n=3 per structure). The automatic contouring was performed across TR ranges of {100, 200, 300, 400, 500, 600, 700, 800, 900, 1000, 1500, 2000, 2500, 3000, 3500, 4000, 4500, 5000} ms and TE ranges of {5, 10, 15, 20, 25, 30, 35, 40, 60, 80, 100, 120, 140, 160, 180, 200} ms. The contrast weighting regions were defined as T1 (TR < 1000 ms and TE ≤ 40 ms), T2 (TR ≥ 1000 ms and TE > 40 ms), and PD (TR ≥ 1000 ms and TE ≤ 40 ms) in agreement with the SyMRI software. The mixed contrast region (TR < 1000 ms and TE > 40 ms) was not evaluated due to its limited use clinically leading to a total of 18 TR points * 16 TE points * 3/4 regions utilized = 216 synthetic contrast-weighted images.

### 2.4. Statistical Analysis

All relevant analysis concerning statistical methods are formulated using the guidelines for reporting Statistical Analyses and Methods in Published Literature (SAMPL)^28^. The DSC^29,30^ and 95^th^ percentile HD (HD95)^31^ were chosen as quantitative measures of contour agreement. For each contrast weighting region, the mean, minimum, and maximum measurement was calculated and reported to sufficiently describe the range of model performance across the region. All calculations were performed in Python 3.7.9 and are publicly available on GitHub via: https://github.com/Lucas-Mc/SyntheticMR_DL-sensitivity. A summary of the data collection and image processing is shown in **Figure 1**.

**Figure 1:**
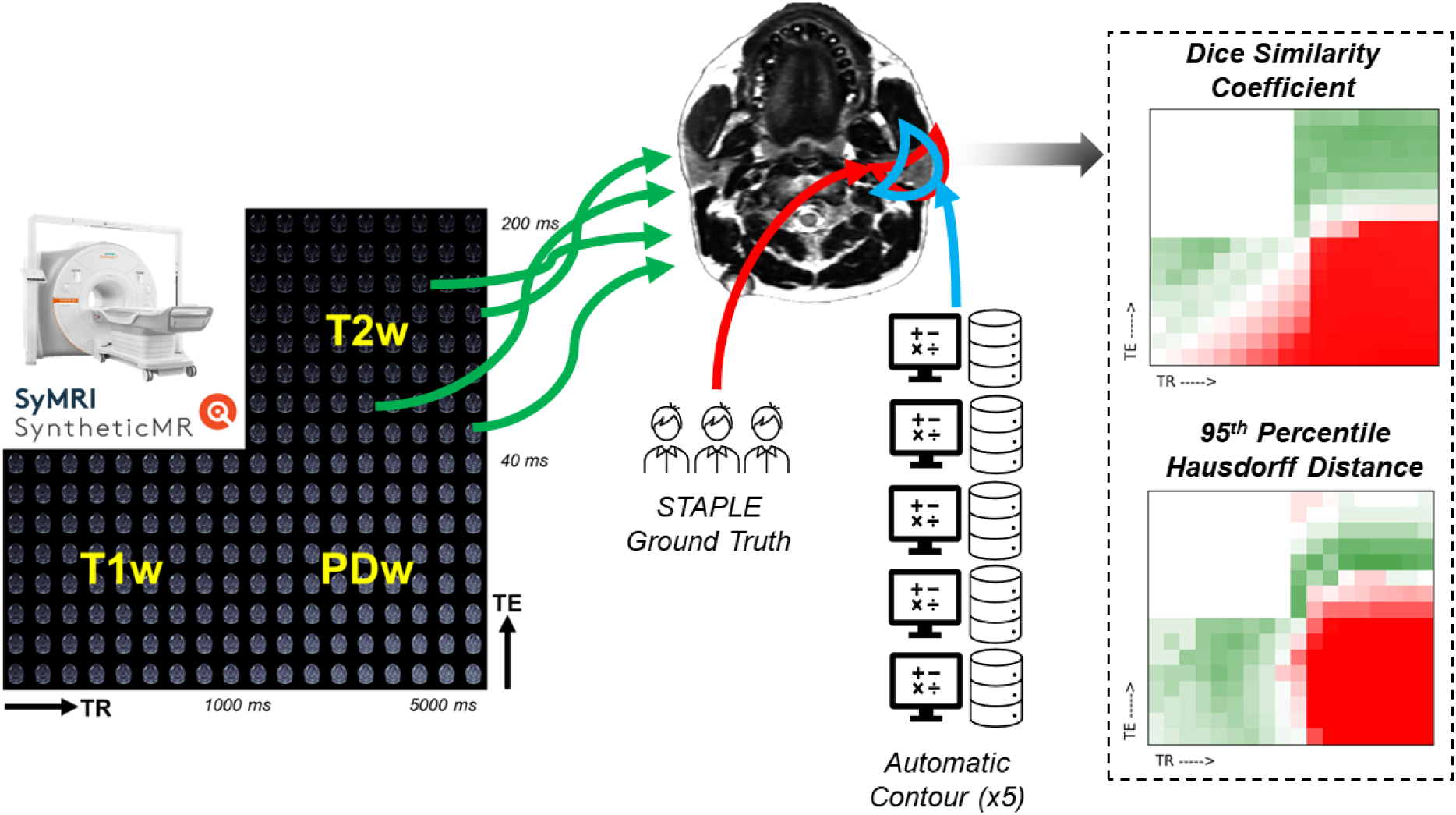
Graphical abstract of the study design. SyntheticMR images are initially acquired on the MR-Simulation device. The raw images are then processed through SyMRI to generate 216 unique contrast weightings of varying TR and TE equally binned across the T1-weighted, T2-weighted, and PD-weighted spectrum. A strong T2-weighted image is used to construct a STAPLE ground truth contour from three medical experts. In parallel, each of the 216 synthetic contrast weighted images are fed into five different automatic contouring algorithms specialized for the salivary glands on T2-weighted MRIs. The agreement between the automatically generated contours and the STAPLE ground truth is measured using the DSC and HD95 and then plotted against the contrast weighting to visually assess algorithm sensitivity.

## 3. Results

### 3.1. Healthy Volunteer Results

**Figure 1** and **Figure 2** demonstrate the automatic contouring model performance for the healthy volunteer quantified using the DSC and HD95, respectively. Further, the plots were made to show the relative TR and TE difference from the acquisition combination closest to the model’s training cohort to demonstrate the amount of flexibility in scan parameters to maintain a desired level of performance.

**Figure 2:**
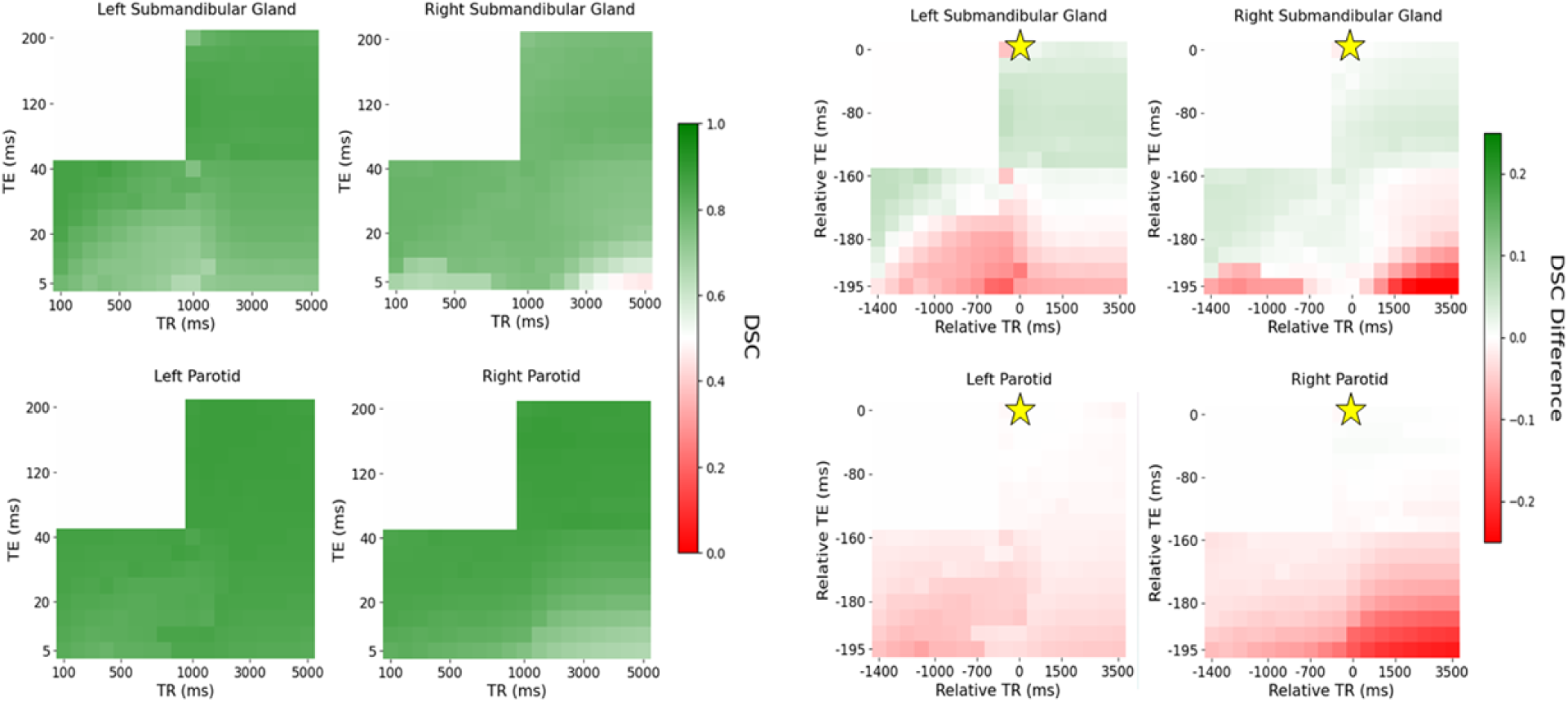
Distribution of DSC (left) and differences in DSC with relative TR and TE compared to the acquisition combination closest to the model’s training cohort (right) across each contrast weighting (T1: bottom left, PD: bottom right, T2: top right for each subfigure, respectively) for the healthy volunteer. The golden star represents the acquisition parameters of the training cohort of TR = 1535 ms and TE = 212 ms.

**Figure 2** and **Figure 3** visually show patches of relatively poor performance of the automatic contouring model from both the DSC and HD95 such as in the bottom right corner of both the T1-weighted and PD-weighted contrast region. Relatively uniform performance is seen in the T2-weighted contrast region; however, some variation is still noted. Quantitative summaries of **Figure 2** and **Figure 3** are given in **Table 2**.

**Figure 3:**
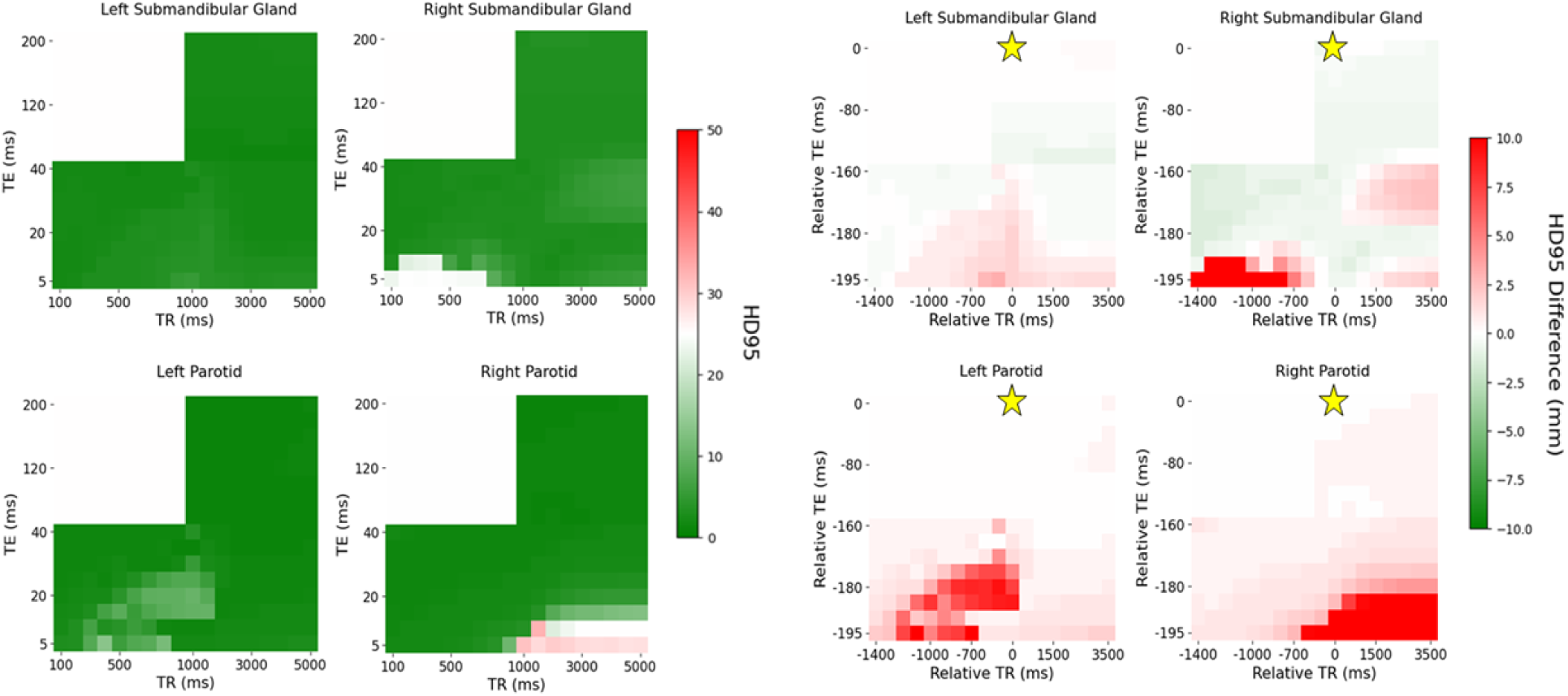
Distribution of HD95 (left) and differences in HD95 with relative TR and TE compared to the acquisition combination closest to the model’s training cohort (right) across each contrast weighting (T1: bottom left, PD: bottom right, T2: top right for each subfigure, respectively) for the healthy volunteer. The golden star represents the acquisition parameters of the training cohort of TR = 1535 ms and TE = 212 ms.

**Table 2:** Distribution of DSC and HD95 for the healthy volunteer for each structure across the T1-, T2-, and PD-weighted contrast ranges.

|  | *Mean [Min, Max]* |  |  |
| --- | --- | --- | --- |
|  | T1-Weighted | T2-Weighted | PD-Weighted |
| **DSC** |  |  |  |
| Left Parotid | 0.84 [0.79, 0.87] | 0.88 [0.87, 0.88] | 0.85 [0.83, 0.87] |
| Right Parotid | 0.85 [0.76, 0.86] | 0.88 [0.86, 0.88] | 0.80 [0.67, 0.87] |
| Left Submandibular | 0.80 [0.66, 0.87] | 0.84 [0.74, 0.86] | 0.78 [0.65, 0.82] |
| Right Submandibular | 0.77 [0.64, 0.80] | 0.78 [0.74, 0.79] | 0.74 [0.45, 0.79] |
| **HD95 (mm)** |  |  |  |
| Left Parotid | 2.00 [1.00, 13.64] | 1.00 [1.00, 1.41] | 1.41 [1.00, 10.34] |
| Right Parotid | 1.73 [1.41, 10.82] | 1.41 [1.00, 1.41] | 3.00 [1.41, 32.29] |
| Left Submandibular | 2.24 [1.73, 5.00] | 2.12 [1.41, 2.45] | 2.45 [2.00, 5.39] |
| Right Submandibular | 2.83 [2.24, 24.92] | 3.16 [3.00, 3.61] | 3.61 [2.45, 6.08] |

Overall, the T2-weighted contrast region exhibited the highest DSC and lowest HD95 compared to the T1- and PD-weighted contrast regions. However, in the right submandibular gland, the T1-weighted contrast region provided the lowest mean HD95 of 2.83 mm compared to 3.16 mm for the T2-weighted contrast region. Additionally, the range of the DSC and HD95 was the lowest in the T2-weighted contrast region, particularly for the HD95 which produced wide ranges outside the region. Quantitatively, the range in the DSC in the T2-weighted region was 0.01, 0.02, 0.12, and 0.05 in the left parotid gland, right parotid gland, left submandibular gland, and right submandibular gland, respectively. The range in the HD95 in the T2-weighted region was 0.41, 0.41, 1.04, and 0.61 mm in the left parotid gland, right parotid gland, left submandibular gland, and right submandibular gland, respectively.

The add more clinical relevance, the DSC plots were threshold by the interobserver variability (IOV) from a study by McDonald at al. with 0.83, 0.84, 0.75, and 0.78 as the cutoff for the left parotid gland, right parotid gland, left submandibular gland, and right submandibular gland, respectively^7^. Similarly, since the analysis from McDonald et al. did not include HD95, the IOV threshold from van der Veen et al. with 4.9, 5.1, 3.1, and 3.1 mm was used as the HD95 cutoff for the left parotid gland, right parotid gland, left submandibular gland, and right submandibular gland, respectively^32^ as shown in **Figure 4**.

**Figure 4:**
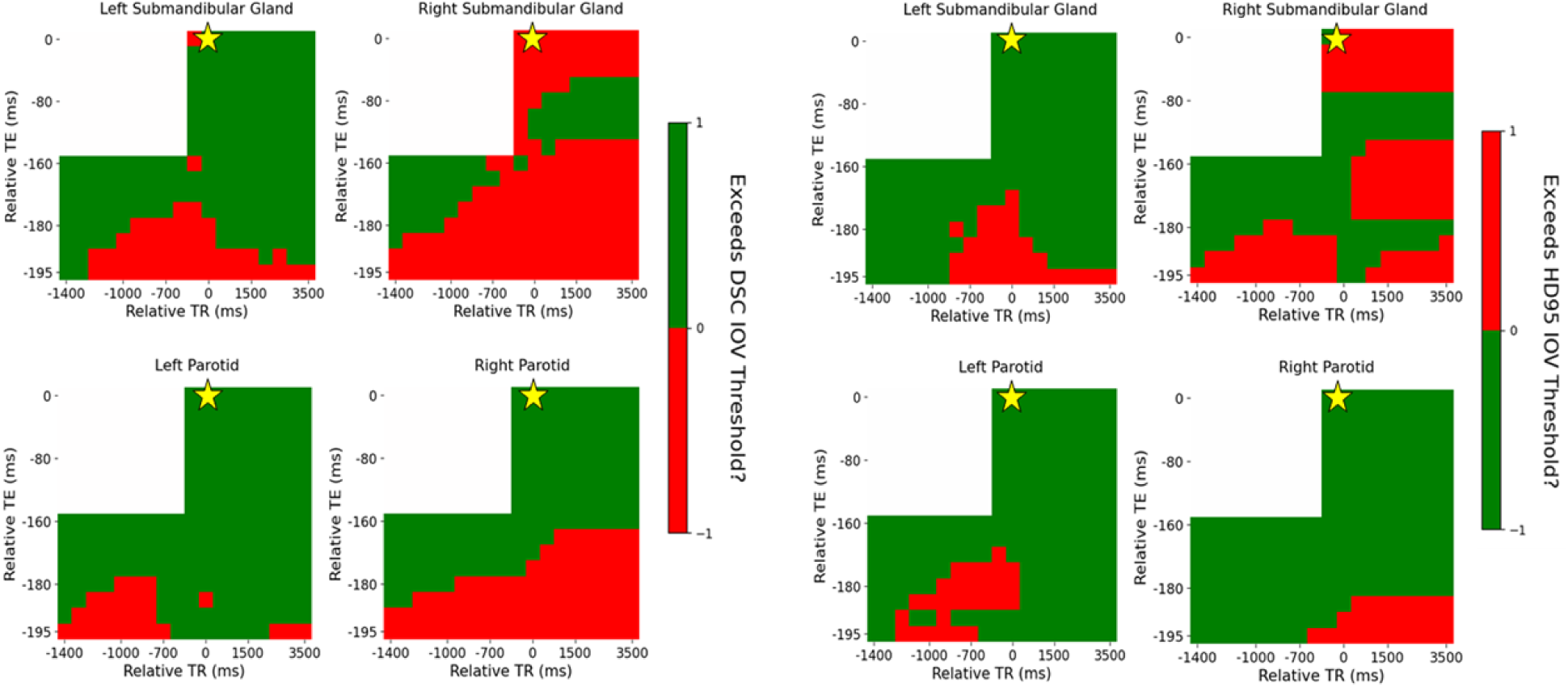
Distribution of DSC (left) and HD95 (right) across each contrast weighting (T1: bottom left, PD: bottom right, T2: top right) with relative TR and TE compared to the acquisition combination closest to the model’s training cohort for the healthy volunteer threshold by the IOV. The golden star represents the acquisition parameters of the training cohort of TR = 1535 ms and TE = 212 ms.

The variation in the IOV across the acquisition parameter combinations is similar between the DSC and HD95. Quantitatively, 88%, 60%, 79%, and 28% of acquisition parameter combinations exceed the IOV threshold for DSC compared to 87%, 88%, 85%, and 50% for HD95 for the left parotid, right parotid, left submandibular, and right submandibular gland, respectively. In some cases, the entire T2-weighted contrast region does not exceed the IOV threshold even though the automatic contouring model was trained on T2-weighted images, specifically in the submandibular glands. Specifically, 99%, and 40% for the DSC and 100%, and 43% for the HD95 in the left submandibular and right submandibular gland, respectively.

### 3.2. Patient Results

**Figure 5** and **Figure 6** demonstrate the automatic contouring model performance for the patient quantified using the DSC and HD95, respectively. Further, the plots were made to show the relative TR and TE difference from the acquisition combination closest to the model’s training cohort to demonstrate the amount of flexibility in scan parameters to maintain a desired level of performance.

**Figure 5:**
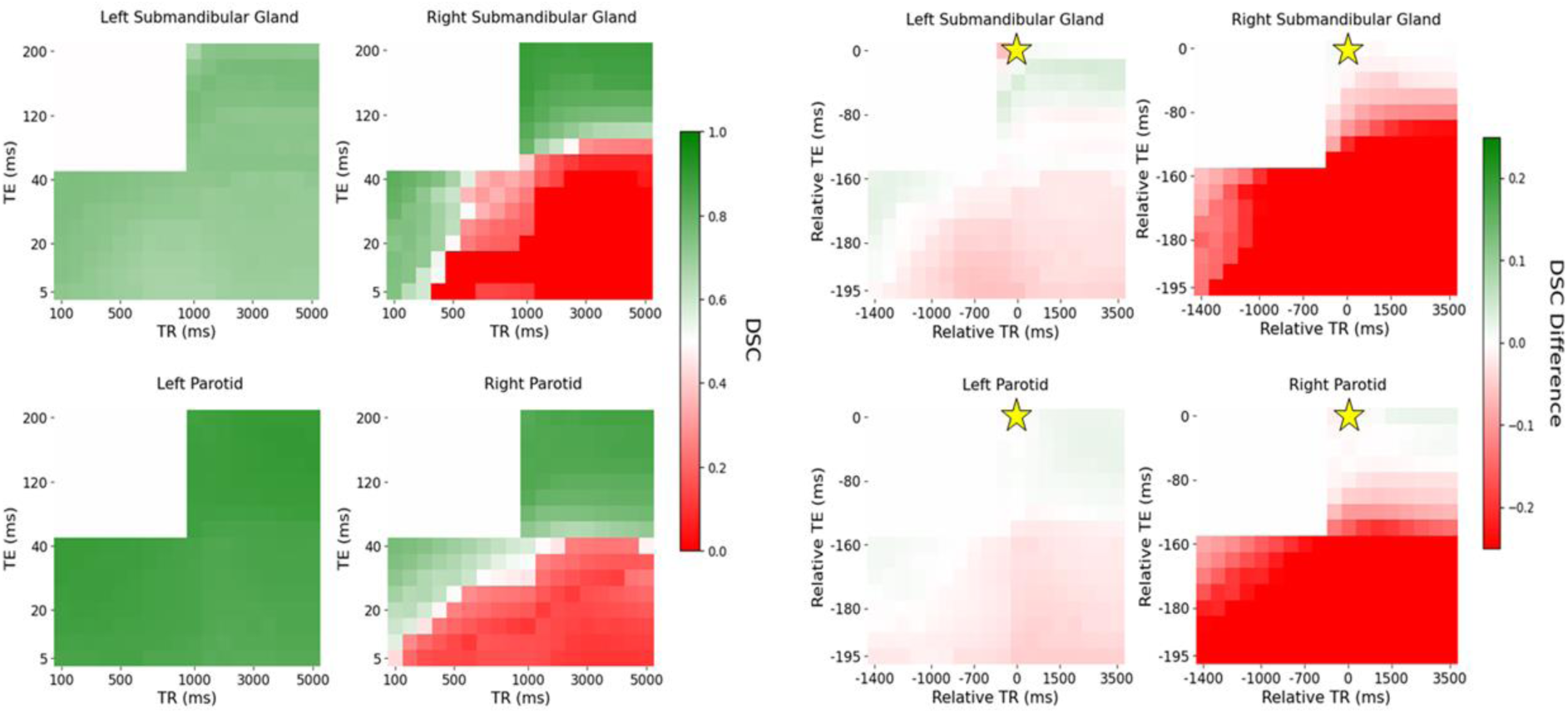
Distribution of DSC (left) and differences in DSC with relative TR and TE compared to the acquisition combination closest to the model’s training cohort (right) across each contrast weighting (T1: bottom left, PD: bottom right, T2: top right for each subfigure, respectively) for the patient. The golden star represents the acquisition parameters of the training cohort of TR = 1535 ms and TE = 212 ms.

**Figure 6:**
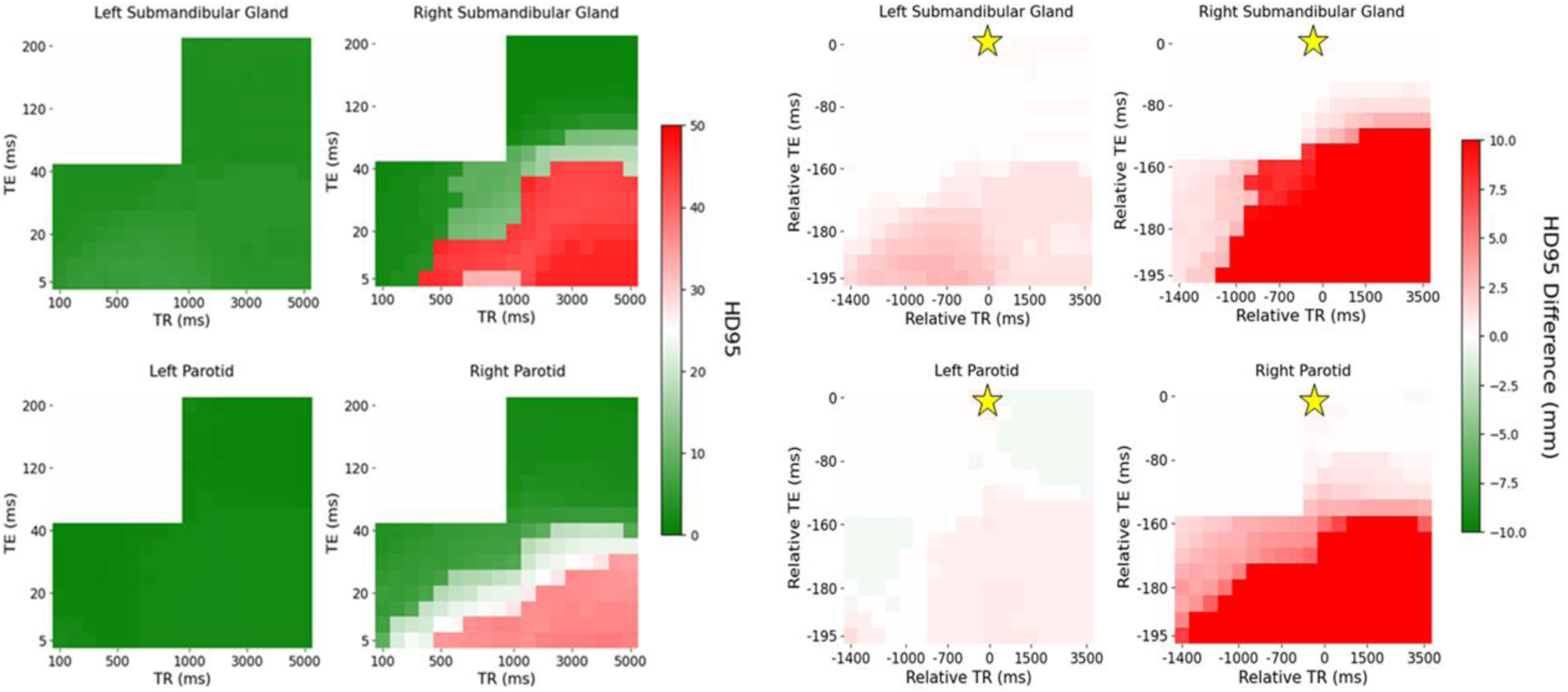
Distribution of HD95 (left) and differences in HD95 with relative TR and TE compared to the acquisition combination closest to the model’s training cohort (right) across each contrast weighting (T1: bottom left, PD: bottom right, T2: top right for each subfigure, respectively) for the patient. The golden star represents the acquisition parameters of the training cohort of TR = 1535 ms and TE = 212 ms.

**Figure 5** and **Figure 6** visually show that the worst model performance was in the PD-weighted range followed by the T1-weighted range and then the T2-weighted range with the highest performance. However, even within the same contrast region, there are visibly large discrepancies in model performance across the range of TR and TE combinations. This is shown for even the T2-weighted range which was the contrast region used for model training.

The pattern of poor performance, similarly, does not follow the same patterns across structures with the right parotid and submandibular gland performing poorly in the PD-weighted range and the respective left side performing comparable to the T2-weighted range. Quantitative summaries of **Figure 5** and **Figure 6** are given in **Table 3**.

**Table 3:** Distribution of DSC and HD95 for the patient for each structure across the T1-, T2-, and PD-weighted contrast ranges. *The model was trained on T2-weighted MRI images.

|  | *Median [Min, Max]* |  |  |
| --- | --- | --- | --- |
|  | T1-Weighted | T2-Weighted* | PD-Weighted |
| **DSC** |  |  |  |
| Left Parotid | 0.87 [0.84, 0.89] | 0.88 [0.85, 0.90] | 0.84 [0.83, 0.86] |
| Right Parotid | 0.49 [0.11, 0.76] | 0.83 [0.65, 0.87] | 0.16 [0.11, 0.57] |
| Left Submandibular | 0.72 [0.67, 0.75] | 0.73 [0.67, 0.77] | 0.70 [0.67, 0.73] |
| Right Submandibular | 0.51 [0.00, 0.82] | 0.81 [0.07, 0.88] | 0.00 [0.00, 0.30] |
| **HD95 (mm)** |  |  |  |
| Left Parotid | 1.41 [1.00, 2.71] | 1.00 [1.00, 2.00] | 2.00 [2.00, 2.24] |
| Right Parotid | 6.85 [3.46, 36.74] | 2.24 [1.73, 5.39] | 35.39 [5.83, 37.85] |
| Left Submandibular | 4.04 [2.83, 5.92] | 3.00 [2.83, 3.32] | 4.12 [3.32, 5.48] |
| Right Submandibular | 3.61 [1.73, 46.08] | 1.45 [1.00, 17.92] | 43.42 [9.70, 46.44] |

Overall, the T2-weighted contrast region exhibited the highest DSC and lowest HD95 compared to the T1- and PD-weighted contrast regions. Additionally, the range of the DSC and HD95 was the lowest in the T2-weighted contrast region, particularly for the HD95 which produced wide ranges outside the region for the right parotid and right submandibular gland. Conversely, the PD-weighted contrast region showed the lowest range in DSC, however its mean score was 0.00 compared to 0.81 for the T2-weighted contrast region. Similarly, in the left parotid, the range in HD95 in the PD-weighted contrast region was 0.24 mm compared to 1.00 mm for the T2-weighted contrast region, however its mean distances were 2.00 and 1.00, respectively. Quantitatively, the range in the DSC in the T2-weighted region was 0.05, 0.22, 0.10, and 0.81 in the left parotid gland, right parotid gland, left submandibular gland, and right submandibular gland, respectively. The range in the HD95 in the T2-weighted region was 1.00, 3.66, 0.49, and 16.92 mm in the left parotid gland, right parotid gland, left submandibular gland, and right submandibular gland, respectively.

Similar to the healthy volunteer, the DSC plots were threshold by the interobserver variability (IOV) for the patient as well as shown in **Figure 7**.

**Figure 7:**
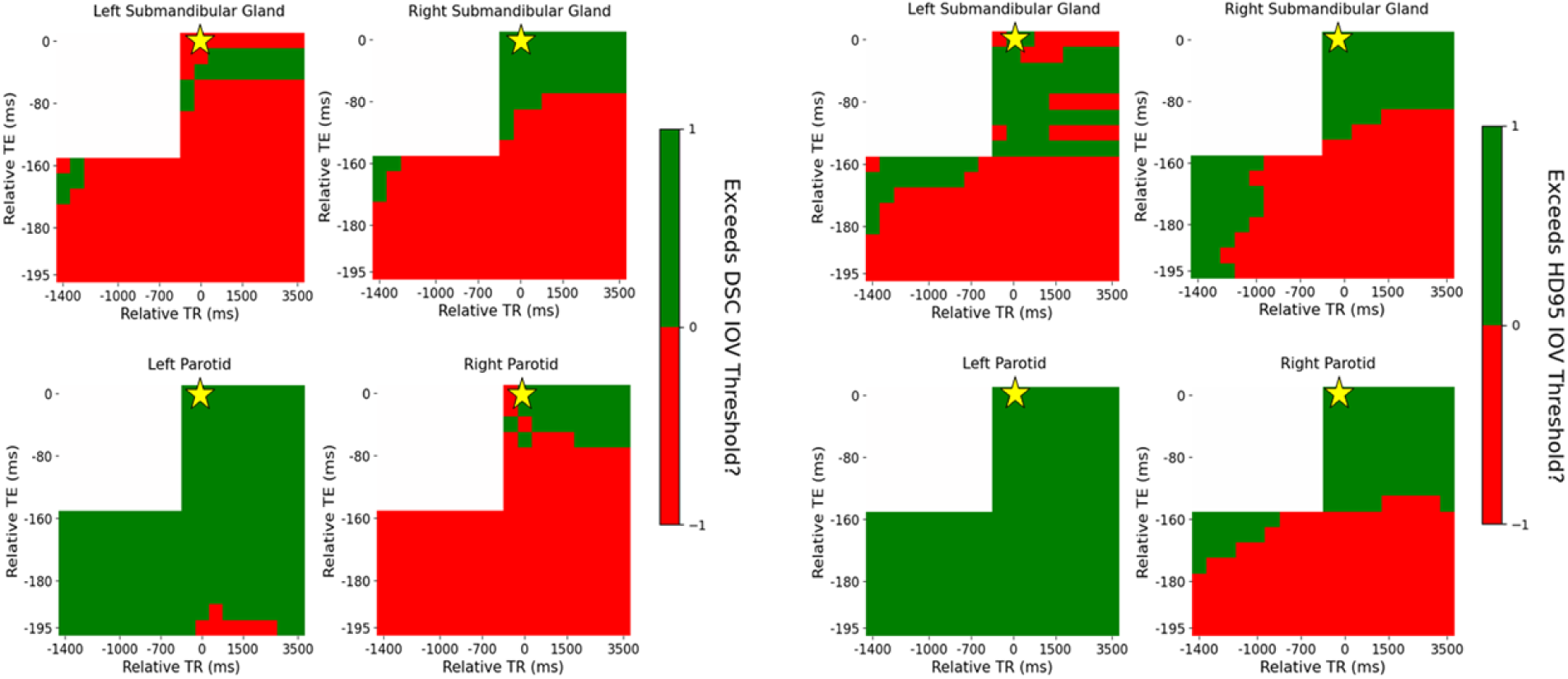
Distribution of DSC (left) and HD95 (right) across each contrast weighting (T1: bottom left, PD: bottom right, T2: top right) with relative TR and TE compared to the acquisition combination closest to the model’s training cohort for the patient threshold by the IOV. The golden star represents the acquisition parameters of the training cohort of TR = 1535 ms and TE = 212 ms.

The variation in the IOV across the acquisition parameter combinations is similar between the DSC and HD95. Quantitatively, 97%, 13%, 10%, and 21% of acquisition parameter combinations exceed the IOV threshold for DSC compared to 100%, 38%, 32%, and 38% for HD95 for the left parotid, right parotid, left submandibular, and right submandibular gland, respectively. The entire T2-weighted contrast region does not exceed the IOV threshold even though the automatic contouring model was trained on T2-weighted images. Specifically, 100%, 40%, 24%, and 57% for the DSC and 100%, 94%, 71%, and 71% for the HD95 in the left parotid, right parotid, left submandibular, and right submandibular gland, respectively.

## 4. Discussion

This study highlights the discrepancy in both DSC and HD95 that can occur across different acquisition parameter combinations (i.e., TR and TE) even within the same contrast weighting region (i.e., T2) due to the slight differences in emphasized T1 and T2 contrast enhancement. Specifically, this study tested the performance of an automatic contouring model trained on T2-weighted MRI images across varying levels of contrast and found that, although the T2-weighted contrast region still exhibited the best results in terms of DSC and HD95, significant discrepancies quantified through the IOV still existed throughout the region. In some cases, the automatic contouring model performed as well on both T1- and PD-weighted images as the T2-weighted images even though the model was trained on T2-weighted images. This confirms that features learned from the T2-weighted images are carried over to the T1-weighted images at certain contrast weighting parameter combinations^33^. This finding corroborates the usefulness of conducting model sensitivity studies to find possible novel and unexpected avenues for transfer learning approaches^34^.

In the healthy volunteer, the minimal difference in DSC in the larger parotid glands (maximum 0.02), but larger difference in the smaller submandibular glands (maximum 0.22). From this observation, care should be taken to comprehensively characterize the model’s performance across acquisition parameters, especially when smaller structures are predicted. Interestingly, comparable performance across the contrast weightings was seen, even with some acquisition parameters in the T1- and PD-weighted contrast regions outperforming those across the T2-weighted contrast region. In the patient, the general range of discrepancy in DSC, even across the T2-weighted contrast region, can be up to 0.2 ignoring the minimum performance of 0.07 in the right submandibular gland, however including that would introduce a range of 0.81. Another interesting finding is that, for some acquisition parameter combinations, comparable DSC to the T2-weighted contrast region is seen in the T1-weighted contrast region. In all structures except the right parotid gland, the maximum DSC in the T1-weighted contrast region exceeds the median DSC in the T2-weighted contrast region. In the left parotid and left submandibular gland, the range of DSC is also comparable between the T1-wegithed contrast region and the T2-weighted contrast region. Interestingly, in the healthy volunteer in the right submandibular gland, a smaller median HD95 was seen in the T1-weighted contrast region compared to the T2-weighted contrast region. Respectively, in the patient, the general range of discrepancy in HD95 even across the T2-weighted contrast region can be up to 3.66 mm ignoring the maximum performance of 17.92 mm in the right submandibular gland, however including that would introduce a range of 16.92 mm. In both the healthy volunteer and the patient, the IOV in the HD95 region is more lenient than the DSC as seen by the increased amount of acquisition parameter combinations which exceed the IOV threshold. Although the analysis here was split by specific anatomical location (i.e., left parotid vs. right submandibular), the ultimate clinical decision should be made at an aggregate level where performance across desired anatomical sites is combined then compared across the different acquisition parameters inside each contrast-weighted region. It is suspected that setup differences are the cause of the performance variation across the lateral direction in the patient with visible B1+ inhomogeneities due to inconsistent head and neck radiofrequency coil placement which aren’t entirely accounted for in the SyMRI post-processing. However, these setup variations should be incorporated and accounted for upon model deployment within a clinical setting since they are within the standard range of expected scenarios.

SyntheticMR’s synthetic T1-, T2-, and PD-weighted images have already been validated in a multi-reader study across 109 patients who determined that the overall diagnostic quality of SyntheticMR images was noninferior to conventional based on a Likert scale (p < 0.001)^35^. This confirms the acceptability of SyntheticMR images as a replacement for conventional approaches. As a more detailed support, a total of 216 synthetic contrast-weighted images would, at best, require around 532 minutes or 8 hours and 52 minutes of scan time using conventional serial scanning approaches^36,37^, however it was achieved using SyntheticMR using a single 4 minute acquisition and approximately 18 minutes of post-processing time (∼30 seconds per 6 images in batch processing, as limited by the SyMRI graphical user interface).

However, it is uncertain whether the quantitative information changes could affect model performance, thus limiting the significance of this study if true. Further, it is unclear if a specific set of acquisition parameters (i.e., the ones used for model training) for a different sequence on a different scanner result in the best matched synthetic images generated from SyMRI which could be the source of seemingly optimal model performance away from the training acquisition parameters in some of the DSC and HD95 difference results. Therefore, a level of calibration should be performed to ensure the synthetic images are sufficiently similar to clinically acquired images at the same acquisition parameters. This can be pursued in future studies which quantitatively compare the model performance among both conventional and SyntheticMR acquisitions at the same TR and TE combinations. Further, the nearest TR and TE studied here was 1500 ms and 200 ms, respectively, which prevented a direct comparison to the TR and equivalent TE used in model training at 1535 ms and 212 ms, respectively, although there should be minimal measurable difference between the two contrasts. Ideally, the human expert contours should be generated on each acquisition parameter combination when comparing to the automatic contouring model output, however this is time- and resource-intensive at the current time, however it is unknown if this would affect human predicted contours significantly compared to the itra-/interobserver variability. Although vague assumptions can be made about different contrast in some organs across the T1-, T2-, and PD-weighted contrast regions^38^, this work has not been investigated rigorously to the authors knowledge and should be addressed in future studies.

A tradeoff must be made when training automatic contouring models between exposing the model to multiple different variations of T2-weighting and maintaining consistency with the same TR and TE for each training image. Often, it may be difficult to collect images of the same TR and TE. One study consisting of 74 prostate cancer patients varied the TR between 2700 – 7291 ms and TE between 100 – 259 ms^39^ while another study for the foot varied the TR between 450 – 652 ms and TE between 12 – 16 ms even across only 10 patients^40^. The same pattern holds in the head and neck with a study including 136 patients across a TR of between 477 – 588 ms and TE of 12 ms for T1-weighted images and TR between 3230 – 5300 ms and TE between 107 – 109 ms for T2-weighted images^41^. These differences may assist the automatic contouring model during training by exposing it to new unseen information, however it will ultimately hinder its performance on the research or clinical deployment phase due to the wide range of possible contrast weightings it may see even though all will be labeled as either T1-weighted or T2-weighted. Several authors have embraced the heterogeneity and actively included multiple acquisition parameters^42–46^ and even contrast weightings^47^ to boost the comprehensive accuracy of their models. Therefore, sensitivity studies similar to this one must be conducted to accurately predict and adjust for the success of the model upon deployment in any setting, thus maximizing model OOD performance.

## 5. Conclusion

Automatic contouring algorithms are seeing increased use in clinical practice due to their efficiency and performance gain compared to traditional human experts. This is especially true in the field of radiation oncology where a large component of daily activities is contouring on MRI images. Currently, these algorithms do not explicitly state their limitations across MRI images of the same contrast (i.e., T2-weighting) even though differences may exist across scanners and institutions. Furthermore, our work indicates that simply stating that an automatic contouring model requires T2-weighted images is not sufficient due to the wide range of performance across T2-weightings. Even more striking is that, for some models, other MRI contrast weightings such as T1 perform equally or even better. For this reason, future work must be done to evaluate automatic contouring algorithms more rigorously across varying MRI contrasts to better understand their sensitivity before deployment into daily clinical practice.

## Contributor Roles Taxonomy (CRediT) Attribution Statement

*Conceptualization:* L.M.; *Data curation:* L.M., Z.B., W.F., A.M.S.A., and K.H.; *Formal analysis:* L.M.; *Funding acquisition:* C.D.F.; *Investigation:* L.M., N.W., S.J.M., and Y.D.; *Methodology:* L.M., K.A.W., and D.T.F.; *Project administration:* C.D.F.; *Resources:* L.M., J.X., D.T., J.S., N.O., K.H. and C.D.F.; *Software:* L.M.; *Supervision:* K.H. and C.D.F.; *Validation:* L.M., N.W., and S.J.M.; *Visualization:* L.M.; *Writing – original draft:* L.M.; *Writing - review & editing:* L.M., Z.B., W.F., A.M.S.A, N.W., S.M., Y.D., J.X., D.T., J.S., N.O., K.A.W., D.T.F., K.H., and C.D.F.

## IRB Statement

All participants provided written informed consent. Volunteers were consented to an internal volunteer imaging protocol (PA15-0418), both approved by the institutional review board at The University of Texas MD Anderson Cancer Center.

## Data Availability Statement

All relevant anonymized image and segmentation files will be made publicly available at the following URL: https://doi.org/10.6084/m9.figshare.28139837. The accompanying code for image visualization and statistical analysis will be made publicly available at the following URL: https://github.com/Lucas-Mc/SyntheticMR_DL-sensitivity.

## Acknowledgements

The authors would like to thank SyntheticMR AB for their assistance in the formal data analysis through their software, SyMRI.

